# The landscape of paediatric infectious disease exposure in a rural sub-Saharan Africa setting in Kilifi, Kenya: longitudinal serological analysis over two decades and priorities for future vaccine development

**DOI:** 10.1101/2024.01.10.24300883

**Authors:** Deirdre F Foley, Timothy K Chege, Joyce Kabagenyi, Karen McCarthy, Elijah T Gicheru, Nelson Kibinge, Angela W Maina, Jacqueline M Waeni, Ralf Clemens, Sue-Ann Costa Clemens, James Tuju, Charles J Sande

**Affiliations:** KEMRI-Wellcome Trust Research Programme, Kilifi, Kenya; Department of Continuing Education, University of Oxford, Oxford, UK; Children’s Health Ireland – Crumlin, Dublin, Ireland; Institute of Global Health, University of Siena, Siena, Italy; International Vaccine Institute, Seoul, Republic of Korea; Oxford Vaccine Group, University of Oxford, Oxford, UK; Nuffield Department of Medicine, University of Oxford, Oxford, UK

## Abstract

**Background:** The paucity of data on the contemporary causes of serious infection among the world’s most vulnerable children means the landscape of emerging paediatric infectious disease remains largely undefined and out of focus on the global vaccine research and development agenda.

**Methods:** We aimed to partially define the paediatric infectious disease landscape in a typical low-income setting in sub-Saharan Africa in Kilifi, Kenya by simultaneously estimating antibody prevalence for 38 infectious diseases using a longitudinal birth cohort that was sampled between 2002 and 2008 and a paediatric inpatient cohort that was sampled between 2006 and 2017.

**Findings:** Among the infectious diseases with the highest antibody prevalence in the first year of life were vaccine-preventable diseases such as RSV (57.4%), mumps (31.5%) and influenza H3N2 (37.3%). Antibody prevalence for *Plasmodium falciparum* shifted substantially over time, from 47% in the mid 2000s to 13% approximately 10 years later corresponding to a documented decline in parasite transmission. A high prevalence of antibodies was also observed in the first year of life for infections for which no licenced vaccines are currently available, including norovirus (34.2%), cytomegalovirus (44.7%), EBV (29.3%) and coxsackie B virus (40.7%). The prevalence to antibodies to vaccine antigens in the local immunisation schedule was generally high but varied by antigen.

**Interpretation:** The data show a high and temporally stable infection burden of RSV, mumps and influenza, providing a compelling evidence base to support progress towards the introduction of these vaccines into the local immunization schedule. The high prevalence of norovirus, EBV, CMV and Coxsackie B provide rationale for increased vaccine research and development investment.

**Funding:** This research was funded by the Wellcome Trust (grant no. WT105882MA).

## Introduction

Though the national under-five mortality rate in Kenya and many other low- and middle-income countries has declined by almost two-thirds since 1990, sub-Saharan Africa still reports the highest childhood mortality rate, and targets for the reduction of global childhood mortality remain unmet.^1^ In the first two decades of this century there have been numerous reports of newly emergent infectious diseases that have caused major human outbreaks – including Rift Valley Fever virus,^2^ pandemic and endemic coronaviruses,^3^ Chikungunya virus,^4^ Ebola virus^5^ and Zika virus^6^ virus among others. While some of these outbreaks have attracted global attention and have been the focus of co-ordinated international vaccine development efforts, many other pathogens with more localised transmission phenotypes in low-income countries have attracted less investment in vaccine research and development. These pathogens continue to exert a considerable and often covert morbidity and mortality burden among the world’s poorest populations.^7^

Many infections such as Rift-Valley Fever, West Nile and Usutu virus, among others, occur primarily in sub-Saharan Africa and cause widespread infections with potentially high case fatality rates. No vaccines currently exist for these and similar high-consequence pathogens that are predominantly localised to low-income countries (LICs). In addition to these major infections, there are many other vaccine-preventable diseases circulating among populations in LICs that continue to exert a serious toll on the health and well-being of these populations, but whose priority for introduction to national immunization schedule remains low.

To make the case for both vaccine introduction and increased investment in vaccine research against locally relevant pathogens, estimates of the infection burden attributable to specific pathogens in vulnerable populations is imperative. To assess this burden, we determined population-level antibody prevalence for 38 distinct infectious disease pathogens in young children from a low-income setting in sub-Saharan Africa. Using multi-target serological analysis of two paediatric cohorts in rural Kenya, we define the prevailing infectious disease landscape in young infants and children over the last two decades and propose priorities for vaccine development and introduction.

## Methods

### Study design and participants

This study was conducted in Kilifi County Kenya, a rural community on the Kenyan Coast. We used two study cohorts to measure disease of 38 infectious disease pathogens in Kenyan infants and young children. The first was a longitudinal birth cohort (Kilifi Birth Cohort, ‘KBC’) recruited between 2002 and 2005 from which cord blood was collected at birth, followed by repeated serum sampling at approximate three-month intervals until children were aged about thirty months of age (Supplementary figure 1). The second was an inpatient cohort comprising of paediatric inpatients admitted to Kilifi County Hospital between 2006 and 2017 with a variety of infectious and non-infectious illnesses (Supplementary figure 2). Serum from each child was assayed for antibodies against the same set of infectious disease pathogens. All data are presented using anonymized codes that are delinked from personally identifiable participant data. This study was conducted in accordance to Good Clinical Laboratory practice principles and was approved by the Scientific Ethics Research Unit of the Kenya Medical Research Institute.

### Procedures

Pathogen-specific antibodies were measured using a customized protein microarray designed to quantify serum IgG to a range of common and emerging infectious diseases. The complete set of pathogens, strains and antigens that were tested on this platform are listed in Supplementary table 1. Data generated on the microarray was extensively validated by relating changes in antibody level to known timings of vaccination or natural infection. Increases in antigen-specific antibodies in the birth cohort time series were confirmed to contemporaneously coincide with independent laboratory diagnoses based on PCR or enzyme immunoassays. An example of this validation approach is illustrated in Supplementary figure 3. Microarray slides were produced by the deposition of two 100pl droplets containing 1ug/ml of the protein antigens onto epoxy-coated polished glass slides by non-contact printing using an Aj003S Arrayjet Sprint microarrayer. Each slide was partitioned into 24 identical mini arrays onto which the complete set of antigens to be assayed were identically printed. Printed slides were allowed to dry at ambient temperature and then placed at 4°C for long-term storage. Prior to sample processing, slides were scanned on a Genepix 4300A fluorescent scanner to obtain background slide fluorescence that would later be subtracted from the final microarray data set to correct for non-specific fluorescence. Slides were then loaded onto hybridization cassettes, washed three times with phosphate buffered saline, and incubated with a blocking buffer comprising phosphate buffered saline supplemented with Tween-20 and 5% bovine serum albumin for an hour at 37°C. A one in thirty dilution of human sera was prepared using phosphate buffered saline supplemented with tween-20 and 5% bovine serum albumin. Different serum samples that were pre-processed in this manner were then aliquoted into distinct mini-arrays and incubated at room temperature for three hours. The slides were then washed thrice, after which a one in four hundred dilution of polyclonal goat anti-human IgG conjugated to Alexa Fluor-647 was added to each mini-array and incubated at room temperature for two hours. The slides were then washed three times and rinsed with Milli-Q water, after which they were scanned using a Genepix 4300A scanner at PMT setting 600 and 100% power. The data was exported to R version 3.6.2 for analysis. The design of the microarray chip and the analytical sample workflow are shown in Supplementary figure 4.

### Statistical analysis

In the birth cohort, antigen-specific seroprevalence was determined by calculating the change in antibody level between successive timepoints in the longitudinal time series. The serological definition of an infection was an increase of ≥ 4000 antibody units (expressed as normalized median fluorescence intensities, MFI) between two serial timepoints and sustained in the subsequent two time points. An example of these serologically defined infections is illustrated in Supplementary figure 3 and in our previously published work^8^. Age-specific seroprevalence for different infections was determined by calculating the proportion of children with serologically defined infections at 12 and 24 months and of age. In the inpatient cohort, seroprevalence estimates at 12 months of age were calculated by first partitioning the antibody data into three-monthly age strata and then identifying a seronegative age stratum at the nadir of maternal antibody decline for each antigen. A serological response threshold for each antigen was then defined as the mean antibody level in the seronegative stratum plus three standard deviations. This threshold was applied to calculate antibody prevalence at subsequent time points. Differences in seroprevalence between the birth and inpatient cohorts were measured using the chi-square 2-sample test for equality of proportions. To visualize age-related changes in antibody level against vaccine antigens in the birth cohort, we used linear spline regression modelling. The non-linear relationship between age and antibody titre was characterized by partitioning the predictor variable (age) into six-monthly segments in which the relationship between age and antibody titre was presumed to be linear. Piecewise linear regression models joined at 6-monthly changepoints starting at birth were developed using the ‘stats’ package available in R.

## Results

To estimate the seroprevalence of antibodies against 38 infectious disease pathogens over the first two decades of this century in children in Kilifi, Kenya, we assayed a total of 1,134 serum samples from two cohorts spanning a period of 15 years (January 2002 to June 2017). Local antibody prevalence in the first of these cohorts, the longitudinal Kilifi Birth Cohort (KBC) was estimated using serial samples collected from 124 children who were recruited at birth. This group was recruited between January 2002 and November 2008 and prospectively sampled at three-monthly intervals for approximately 30 months. An average of eight serum samples were collected from each child in the longitudinal series, resulting in a total collection of 958 serum samples (table 1). The second cohort was a paediatric inpatient (IP) cohort in which a single admission sample was collected from 176 children, who ranged between one day and 57 months of age and who had been admitted to hospital with a variety of medical conditions (table 1). A summary of the sampling frame for both the longitudinal and inpatient cohorts is shown in Supplementary figures 1 and 2 respectively.

**Table 1.** Demographics and patient sampling methods of both cohorts within the study. Each participant in the longitudinal birth cohort had cord blood collected at birth, followed by repeated serum sampling at three-monthly intervals. The inpatient cohort (IP) had serum from each child sampled at single time point during a hospital admission and assayed for antibodies against the same set of infectious disease pathogens.

|  | Longitudinal cohort (KBC) | Inpatient cohort (IP) |
| --- | --- | --- |
| Number of children in cohort | 124 | 176 |
| Recruitment dates | 28<sup>th</sup> Jan 2002- 25<sup>th</sup> Nov 2008 | 20<sup>th</sup> Dec 2006 – 10<sup>th</sup> June 2017 |
| Average number of samples per child (range) | 8 (5 – 11) | 1 |
| Median age in months at recruitment (range) | 0 | 7.2 (0-57) |
| Median age at exit from cohort (range) | 23 (8.3 – 30.2) | NA |
| Total serum samples tested | 958 | 176 |

Antibody prevalence for all 38 infectious diseases featured on the protein array was estimated at 12- and 24-months age in the birth cohort as shown in figure 1. Overall seroprevalence ranged from 0% for ebolavirus and yellow fever virus, to 57% for Respiratory Syncytial Virus (RSV) in the first year of life. To facilitate detailed analysis, antibody data were further stratified into four groups as follows: high prevalence infections (defined as seroprevalence ≥ 25%) that were vaccine-preventable, but not currently included on the Kenya Expanded Programme on Immunization (EPI) schedule (i.e. high prevalence, not on EPI or HP-NEPI), high prevalence diseases for which no licensed vaccines exist (high prevalence, no vaccine or HP-NV), low prevalence (LP) infections (seroprevalence <25%) regardless of vaccine availability, and diseases whose vaccines are current included in the Kenya EPI (i.e. Established in EPI or EEPI).

**Figure 1.**
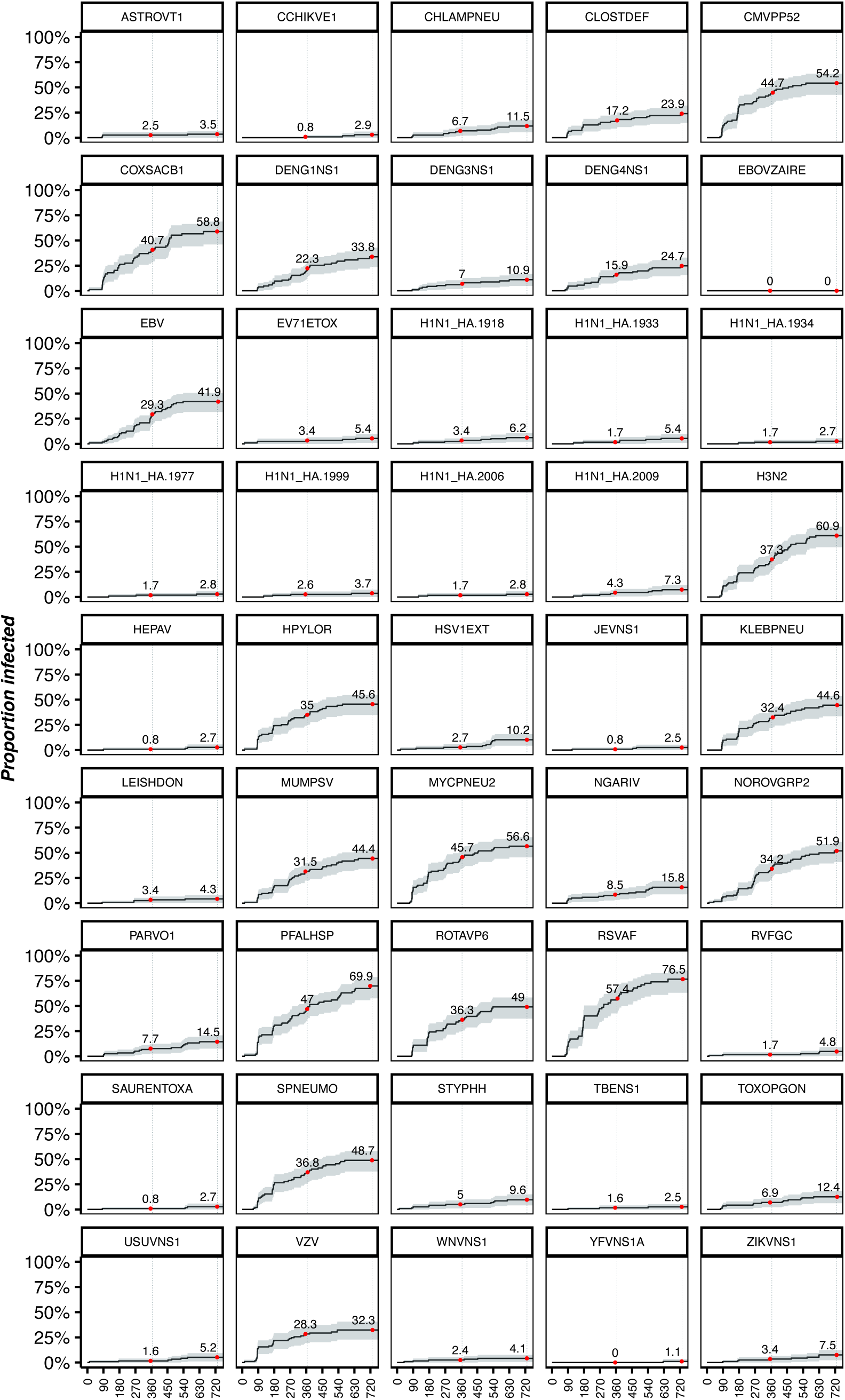
Reverse Kaplan-Meier survival curve demonstrating cumulative antibody prevalence for each target within the panel of infectious diseases pathogens featured on the protein array over a two-year period. The red dots indicate seroprevalence at 12- and 24-months withing the longitudinal cohort. Each curve measures the proportion of population infected (Y-axis) with age in days (X-axis).

RSV, Mumps virus and Influenza virus H3N2 satisfied the HP-NEPI classification criteria in both the longitudinal and inpatient cohorts, with 12-month antibody seroprevalence of 57.4% and 56% respectively for RSV, 32% and 31% respectively for Mumps virus and 37% and 25% respectively for H3N2 influenza. By 24 months of age, RSV seroprevalence had increased to 76.5% in the longitudinal cohort, while Mumps and H3N2 seroprevalence at this time point was 44.4% and 60.9% respectively (table 1 and figure 1). VZV seroprevalence partially satisfied these criteria, with 28% seroprevalence in the longitudinal cohort, later declining to 15% in the inpatient cohort. A similar pattern of antibody prevalence decline was observed for *Plasmodium falciparum*, whose 12-month seroprevalence the longitudinal cohort was 47%, declining to 13% in the inpatient cohort (figure 1 and figure 3). Two additional infections, *Streptococcus pneumoniae* and Rotavirus were initially categorized in the HP-NEPI with estimated 12-month seroprevalence rates of 36% and 45.9% respectively in the longitudinal cohort (figure 1 and table 2) and 18% and 27% in the inpatient cohort (figure 3) but later transitioned to the EEPI category when their respective vaccines were introduced to the local EPI schedule in 2011 and 2014 respectively.

**Table 2.** Table 2 lists the pathogens assayed and the associated 12- and 24-month seroprevalence for each. The clinical sub-group is denoted beside each target. LP=Low Prevalence, HP-NEPI=High Prevalence-National Expanded Programme on Immunization, EEPI=Established Expanded Programme on Immunization, HP-NV=High Prevalence-No Vaccination

| Antigen code | Pathogen | Seroprevalence at 12 months (95% CI) | Seroprevalence at 24 months (95% CI) | Group |
| --- | --- | --- | --- | --- |
| EBOVZAIRE | Ebola virus – Zaire strain | 0 (0-0) | 0 (0-0) | LP |
| YFVNS1A | Yellow fever virus | 0 (0-0) | 1.1 (0-3.2) | LP |
| JEVNS1 | Japanese Encephalitis virus | 0.8 (0-2.4) | 2.5 (0-5.3) | LP |
| TBENS1 | Tick-borne encephalitis virus | 1.6 (0-3.9) | 2.5 (0-5.3) | LP |
| HEPAV | Hepatitis A virus | 0.8 (0-2.4) | 2.7 (0-5.6) | LP |
| H1N1_HA.1934 | Influenza A (H1N1 1934) | 1.7 (0-3.9) | 2.7 (0-5.6) | LP |
| SAURENTOXA | Staphylococcus aureus | 0.8 (0-2.4) | 2.7 (0-5.7) | LP |
| H1N1_HA.2006 | Influenza A (H1N1 2006) | 1.7 (0-4.1) | 2.8 (0-5.9) | LP |
| H1N1_HA.1977 | Influenza A (H1N1 1977) | 1.7 (0-4.1) | 2.8 (0-5.9) | LP |
| CCHIKVE1 | Chikungunya virus | 0.8 (0-2.5) | 2.9 (0-6.1) | LP |
| ASTROVT1 | Astrovirus | 2.5 (0-5.3) | 3.5 (0.1-6.8) | LP |
| H1N1_HA.1999 | Influenza A (H1N1 1999) | 2.6 (0-5.5) | 3.7 (0.1-7.2) | LP |
| WNVNS1 | West Nile virus | 2.4 (0-5.1) | 4.1 (0.5-7.6) | LP |
| LEISHDON | Leishmania donovani | 3.4 (0.1-6.6) | 4.3 (0.5-7.9) | LP |
| RVFGC | Rift valley fever virus | 1.7 (0-3.9) | 4.8 (0.6-8.8) | LP |
| USUVNS1 | Usutu virus | 1.6 (0-3.8) | 5.2 (1-9.1) | LP |
| H1N1_HA.1933 | Influenza A (H1N1 1933) | 1.7 (0-4) | 5.4 (1.1-9.5) | LP |
| EV71ETOX | Enterovirus 71 | 3.4 (0.1-6.5) | 5.4 (1.1-9.6) | LP |
| H1N1_HA.1918 | Influenza A (H1N1 1918) | 3.4 (0.1-6.6) | 6.2 (1.6-10.6) | LP |
| H1N1_HA.2009 | Influenza A (H1N1 2009) | 4.3 (0.5-7.9) | 7.3 (2.3-12.1) | LP |
| ZIKVNS1 | Zika virus | 3.4 (0.1-6.6) | 7.5 (2.3-12.4) | LP |
| STYPHH | Salmonella enterica (serovar typhi) | 5 (1-8.8) | 9.6 (4-14.8) | LP |
| HSV1EXT | Herpes simplex virus 1 | 2.7 (0-5.6) | 10.2 (4.3-15.8) | LP |
| DENG3NS1 | Dengue virus Type 3 | 7 (2.2-11.5) | 10.9 (4.9-16.6) | LP |
| CHLAMPNEU | Chlamydia pneumoniae | 6.7 (2.1-11.1) | 11.5 (5.4-17.2) | LP |
| TOXOPGON | Toxoplasma gondii | 6.9 (2.2-11.4) | 12.4 (6.1-18.4) | LP |
| PARVO1 | Parvovirus 1 | 7.7 (2.7-12.4) | 14.5 (7.6-20.8) | LP |
| NGARIV | Ngari virus | 8.5 (3.3-13.4) | 15.8 (8.8-22.2) | LP |
| CLOSTDEF | Clostridium difficile | 17.2 (9.8-23.9) | 23.9 (14.9-31.8) | LP |
| DENG4NS1 | Dengue virus Type 4 | 15.9 (8.7-22.6) | 24.7 (15.5-32.9) | LP |
| VZV | Varicella zoster virus | 28.3 (19.3-36.3) | 32.3 (22.9-40.6) | HP-NEPI |
| DENG1NS1 | Dengue virus Type 1 | 22.3 (13.8-29.9) | 33.8 (23.4-42.8) | LP |
| EBV | Epstein-Barr virus | 29.3 (20.2-37.3) | 41.9 (31.7-50.6) | HP-NV |
| MUMPSV | Mumps virus | 31.5 (22.4-39.5) | 44.4 (34.1-53) | HP-NEPI |
| KLEBPNEU | Klebsiella pneumoniae | 32.4 (22.7-40.9) | 44.6 (33.7-53.7) | HP-NV |
| HPYLOR | Helicobacter pylori | 35 (25.1-43.6) | 45.6 (34.9-54.6) | HP-NV |
| ROTAVP6 | Rotavirus | 36.3 (25.6-45.5) | 49 (37.5-58.4) | HP-NEPI-CEPI* |
| NOROVGRP2 | Norovirus | 34.2 (24.8-42.5) | 51.9 (40.9-60.9) | HP-NV |
| CMVPP52 | Cytomegalovirus | 44.7 (33.5-54) | 54.2 (42.6-63.5) | HP-NV |
| MYCPNEU2 | Mycoplasma pneumoniae | 45.7 (35-54.6) | 56.6 (45.5-65.4) | HP-NV |
| COXSACB1 | Coxsackie B | 40.7 (29.1-50.4) | 58.8 (46-68.7) | HP-NV |
| H3N2 | Influenza A (H3N2 1999) | 37.3 (27-46.2) | 60.9 (49.5-69.8) | HP-NEPI |
| SPNEUMO | Streptococcus pneumoniae | 45.9 (33.8-55.7) | 64.1 (51.8-73.3) | HP-NEPI-CEPI* |
| PFALHSP | Plasmodium falciparum | 47 (35.8-56.2) | 69.9 (57.5-78.6) | HP-NEPI |
| RSVAF | Respiratory syncytial virus | 57.4 (46.9-70.3) | 76.5 (68-88.8) | HP-NV |

The HP-NV group for which no licensed vaccines are currently available was made up of four infectious diseases, whose 12-month seroprevalence estimates were consistently above 25% in both cohorts. Norovirus (34% in both the longitudinal and inpatient cohorts), Cytomegalovirus (44.7% longitudinal cohort, 29% inpatient cohort), Coxsackie B virus (40.7% longitudinal cohort, 25% inpatient cohort) and Epstein-Barr virus (29% in both the longitudinal and inpatient cohorts) are displayed in table 2 and figure 3. For both HP-NEPI and HP-NV groups, these seroprevalence estimates rose substantially at 24 months of age (figure 1, table 2 and figure 3). Several infections including *Klebsiella pneumoniae, Helicobacter pylori* and *Mycoplasma pneumonia* exhibited high antibody seroprevalence above the 25% threshold in the inpatient cohort but lower seroprevalence below this level in the longitudinal cohort (figure 3).

The low prevalence (LP) group featured a variety of viral pathogens as shown in table 2, including flaviviruses such as Dengue viruses, Zika virus, Chikungunya virus, Ebola virus, West-Nile virus, tick-borne encephalitis, and yellow fever virus. Except for dengue, all other flaviviral infections had antibody prevalence below 5% at 12 months of age as shown in table 2. Twelve-month seroprevalence for dengue viruses ranged from 7% for dengue virus type 3 in the longitudinal cohort and 9% in the inpatient cohort, to 22.3% for dengue virus type 1 in the longitudinal cohort and 15% in the inpatient cohort. Among the infections studied in the LP group were seven Influenza H1N1 viruses, including the 1918 and 2009 pandemic virus strains. The seroprevalence of antibodies against all H1N1 Influenza A viruses, including the pandemic strains, was consistently below 15% at 12 months of age in both cohorts (figure 3). Other LP infections included hepatitis A, *Staphylococcus aureus*, Usutu virus, Rift valley fever virus, Astrovirus, Enterovirus 71 virus and *Leishmania donovani,* with antibody prevalence estimates at or below 3% by the end of the first year of life.

The EEPI group included infections whose respective vaccines are currently included in the Kenya EPI and showed consistently high seroprevalences. In the longitudinal cohort, we evaluated the antibody response against three antigenic targets in the pentavalent vaccine formulation and found antibody prevalence for *Corynebacterium diphtheriae*, *Bordetella pertussis* and Hepatitis B virus of 70.8%, 56.6% and 46% respectively at 12 months of age, rising to 80.5%, 66.1% and 53.6% respectively by 24 months of age (figure 2). We also examined temporal changes in antibody level to determine whether the timing of the antibody response to these antigens was consistent with vaccination or natural exposure. For all three antigens, there was a rapid rise in antibody level within the first three months of life, coinciding with the prescribed timing of 3 pentavalent vaccine doses on the local EPI schedule (6 weeks, 10 weeks, and 14 weeks) as seen in figure 2. The peak population-level antibody titre occurred shortly after the final dose, followed by a variable temporal pattern of antibody decay that was observed post peak (figure 2). Pertussis-specific titres remained stable over time, while antibodies targeting the diphtheria toxin and hepatitis B surface antigen decayed at a faster rate during follow-up but remained significantly above cord blood titre levels despite the wane. Comparison of these antibody dynamics with those of unrelated antigens that are not part of the local vaccination schedule (Ebolavirus, EBV and H1N1 influenza (1999) virus) showed that temporal kinetics of these alternative antibody specificities were antigen-specific and did not correspond to the scheduled timing of the three pentavalent vaccine doses (figure 2).

**Figure 2.**
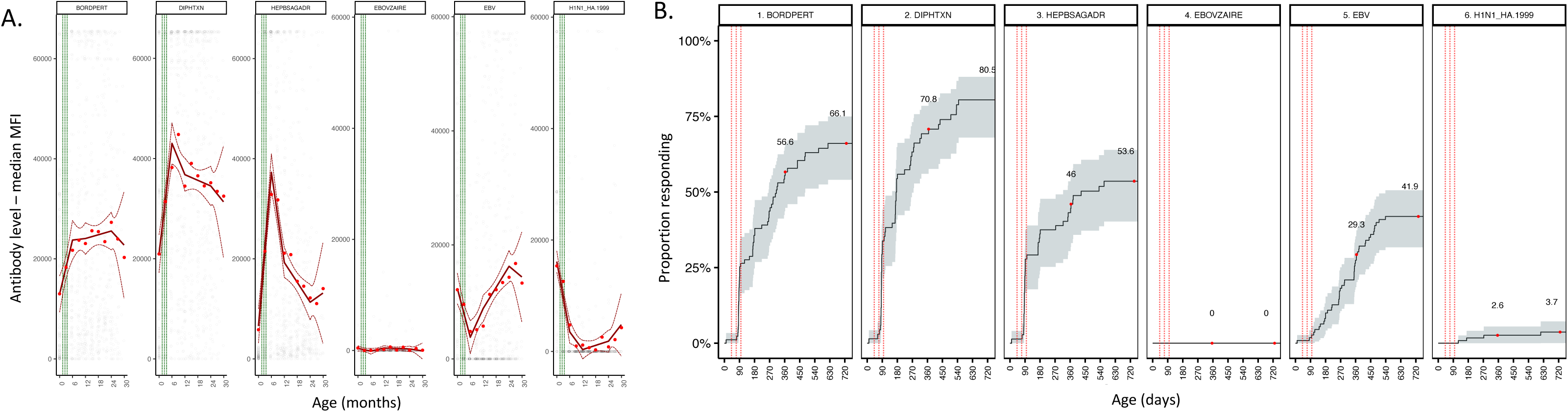
**Figure 2A.** Antibody trends measured in mean fluorescent intensity (MFI) using linear spline modelling. Aise in MFI levels follow immunization or natural infection. An initial fall of MFI after birth represents maternal antibody decay in targets with high population seroprevalence. A later fall in MFI suggests a waning of antibody response. The vertical green lines indicate timepoints of pentavalent vaccination. Dashed red lines represent 95% CI. **Figure 2B.** Reverse Kaplan-Meir plot survival curve demonstrating the cumulative proportion of the cohort responding to immunization or exposure to the same targets as fig. 2A over the first two years of life. Red dots indicate 12- and 24-month timepoints. Dashed red lines indicate pentavalent vaccination.

**Figure 3.**
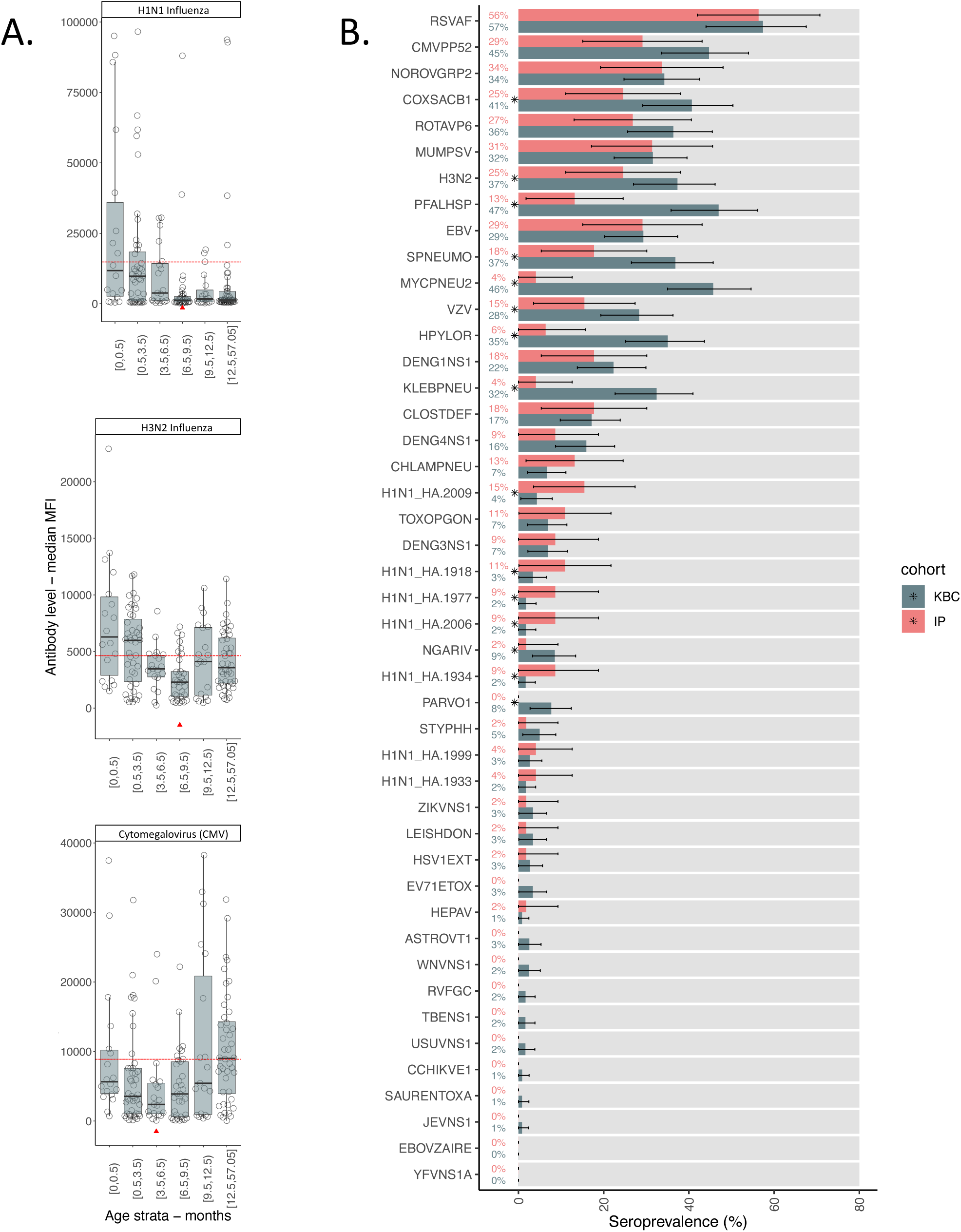
**Figure 3A** – Inpatient cohort antibody response to three targets i) influenza H1N1, ii) influenza H3N2, and iii) CMV. Antibody values are measured using mean fluorescent intensity (MFI). The horizontal axis stratifies patients by age in months, up to five years of age. The red triangles denote the mean of the seronegative group in each figure. The dashed red line denotes seroconversion, ie two standard deviations above the median antibody level of the seronegative stratum. **Figure 3B** – Horizontal bar chart contrasting both the longitudinal and inpatient cohort specific antibody response. (CI 95%). The pathogens are labelled in code available in Supplementary table 1.

## Discussion

The main objective of this study was to describe the contemporary landscape of paediatric infectious disease among children in a rural, sub-Sharan African setting in Kenya by estimating the antibody prevalence for a range of infectious diseases. Our data demonstrate a considerable and sustained local infection burden from several vaccine preventable diseases in the first year of life including RSV, Mumps virus and H3N2 influenza. Just under 60% of infants in both the longitudinal birth cohort and the inpatient cohort had detectable serum antibody against RSV at twelve months of age, rising to around 80% seroprevalence by their second birthday. RSV is the most important cause of severe childhood respiratory illness in the world^9^, with the mortality burden being disproportionately borne by children in low-resource settings^10^ due to poor healthcare infrastructure for the supportive care of critically ill children, including supplemental oxygen and mechanical ventilation. A new maternal vaccine against RSV with a reported efficacy of 82% against severe disease in the first 3 months^11^ of life has recently been approved by the Food and Drug administration (FDA) in the United States^12^ and the European Medicines Agency^13^. While extensive surveillance studies in the United States and Europe have long established the health and economic burden attributable to RSV in those settings, similar data in lower resource settings have been sparse. The data presented here indicates a significant infection burden due to RSV among infants in sub-Saharan Africa and contributes to the evidence base that will inform decisions on whether to introduce newly licensed maternal RSV vaccines in these settings. Our data aligns with longitudinal birth cohort studies in the United States which demonstrated 68% and 82% antibody prevalence against RSV at the end of the first and second years of life respectively.^14^

Around a third of infants in both study cohorts had serologic evidence of mumps infection at 12 months of age rising to about 44% by their second birthday. Mumps is a highly contagious febrile infection, that can lead to potentially debilitating sequelae including meningitis, hydrocephalus, pancreatitis, and hearing loss.^15^ A safe and highly effective vaccine against mumps has been available since the 1960s, initially as a stand-alone vaccine and later as a multivalent formulation comprising measles, mumps, and rubella virus antigens.^16^ Our data indicate a substantial local burden of natural mumps infection in Kenyan children and provide rationale for vaccine introduction in the local paediatric immunization schedule to address this sizeable infection burden.

Our analysis of influenza infection in infants and young children found a high prevalence of antibodies to H3N2 influenza and a low prevalence of H1N1-specific antibodies. We found uniformly low prevalence of antibodies specific to broad range of H1N1 strains, including the 1918 and 2009 pandemic strains as well as seasonal H1N1 strains isolated in 1933, 1934, 1977, 1999 and 2006. These observations align with results from previous studies of African children, where the incidence of seasonal H1N1 influenza infection in infancy was reported to be around 6 per 10,000 infants.^17,18^ We identified a much greater prevalence of antibodies against the H3N2 strain of seasonal influenza, with antibody prevalence remaining above 25% in both study cohorts at 12 months of age. Paediatric influenza vaccines licenced for use in children over six months have been available for several years now. Although many studies in this population have demonstrated a clear benefit of vaccination against severe disease,^19^ vaccine evaluation studies have shown modest vaccine effectiveness of seasonal influenza vaccination in children, particularly against the H3N2 strain of the virus.^20,21^

For some pathogens, we found a shifting pattern of antibody prevalence over time. For example, antibody prevalence for *Plasmodium falciparum* was initially 47% in the longitudinal cohort which was sampled between 2002 and 2008 but later declined to 13% in the inpatient cohort (sampled between 2006 and 2017). This decay in antibody prevalence corresponds with documented evidence of a local decline in malaria disease burden in infants during that period. The proportion of infants admitted to hospital in Kilifi Kenya with a positive *P. falciparum* test declined from over 50% in the early 2000s to less than 10% by 2014.^22^ Our data therefore largely confirm these temporal changes in the local paediatric disease burden attributable to falciparum malaria and indicates that this serological approach would be useful in measuring future changes in disease trends at the population level.

We also detected a change in antibody prevalence for two infections whose vaccines were not part of the local immunization schedule at the time of sampling the longitudinal cohort, but that were later introduced soon after the initiation of inpatient cohort sampling. The decline in prevalence of antibodies against *S. pneumoniae* from 36% in the longitudinal cohort to 18% in the inpatient cohort and from 36.3% to 27% for Rotavirus, corresponds with the introduction of highly effective vaccines for these targets into the local EPI schedule in 2011^23^ and 2014^24^ respectively.

A limitation of our study is that the demographics of the inpatient cohort differ from the birth cohort in that the median age is younger (7.2 months), with a range of 0-5 years. The birth cohort were tracked at regular intervals to 24 months of age mapping seroprevalence trends over the first two years of life. While the inpatient group represents a contemporary glimpse into childhood seroprevalence of infection, direct comparison of data between the cohorts should be interpreted in a conservative manner. Group B streptococcus antibody was not included as a for target for serological analysis within this study and represents a potential future research opportunity.

Our study found a high antibody prevalence for several infections against which no licensed vaccines currently exist. Notable among these were EBV, the cause of infectious mononucleosis^25^ and malignant Burkitt’s lymphoma,^26^ CMV which causes congenital infection associated with severe neurological sequelae including sensorineural hearing loss and cognitive impairment^27,28^ and Coxsackie B virus whose symptoms range from acute febrile manifestations to myocarditis.^29^ In addition to these infections, we found that more than a third of infants in both study cohorts had antibodies against Norovirus, a highly contagious cause of acute severe gastroenteritis. ^25^ Since the introduction of effective rotavirus vaccines, norovirus has become the leading cause of vomiting and diarrhea in many countries around the world.^30^ By establishing the considerable burden of disease attributable to these infections, our data supports the case for increased investment in vaccine research and development as well as broader surveillance in diverse populations, particularly in low resource settings. In contrast to these infections, we found a low prevalence of antibodies to a range of infections, including most flaviviruses, *Staphylococcus aureus*, Enterovirus 71 and *Leishmania donovani*.

Finally, our assessment of the temporal kinetics of antibodies specific to three vaccine antigens in the pentavalent vaccine showed a population-level increase in the titre of antibodies against diphtheria, Hepatitis B and *Bordetella pertussis* that coincided with the prescribed age for pentavalent vaccine dosing at 6, 10 and 14 weeks. The estimated seroprevalence at 12 months was highest for diphtheria at around 71%, followed by pertussis and hepatitis B at 56.7% and 46% respectively. Since these antigens are delivered in a single formulation, the variable antibody prevalence estimates might be explained by several factors including differential background rates natural infection, varying propensities for immune stimulation and differential rates of post-vaccination antibody decay. Even by adopting the most sanguine interpretation of these data, that the estimated seroprevalence of diphtheria (71%) reflects true vaccine coverage, the data indicate that a substantial proportion of infants remain unprotected by the end of infancy, thereby providing a justification for more detailed assessment of suboptimal immunization in children from such settings.

In summary, our research advocates for the urgent introduction of vaccines against RSV, mumps and seasonal influenza into the local immunization schedule. In the case of RSV, maternal vaccination should be paired long-acting immunoprophylactic monoclonal antibodies to provide extended protection to the end of infancy following the decay of maternal antibodies. In conclusion, our study is the first major multi-target sero-surveillance study in African children and the results provide the evidence base to reconsider priorities for the development and introduction of vaccines for a low resource setting in sub-Saharan Africa.

## Data Availability

All data produced in the present study are available upon reasonable request to the authors

## Supplementary figures

**Supplementary figure 1.**
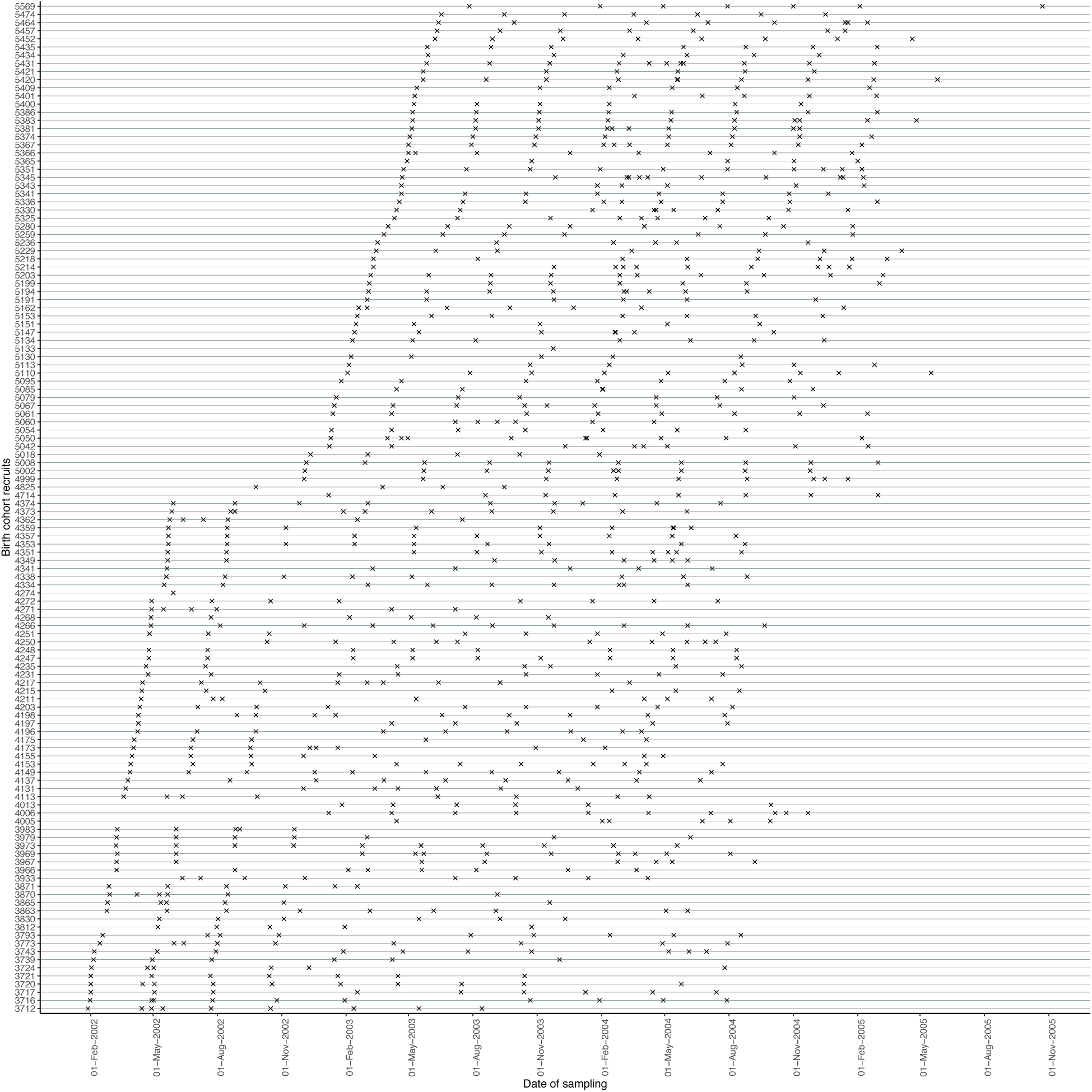
Time points at which data was collected on each individual patient within the longitudinal cohort recruited between 2002 and 2005. The vertical axis uses the study number of each recruit. The first ‘x’ on the horizontal line designated to each recruit represents cord blood sampling at birth, followed by repeated serum sampling at approximate three-month intervals until children were aged about thirty months of age. The y-axis contains anonymized codes that are delinked from personally identifiable participant data.

**Supplementary figure 2.**
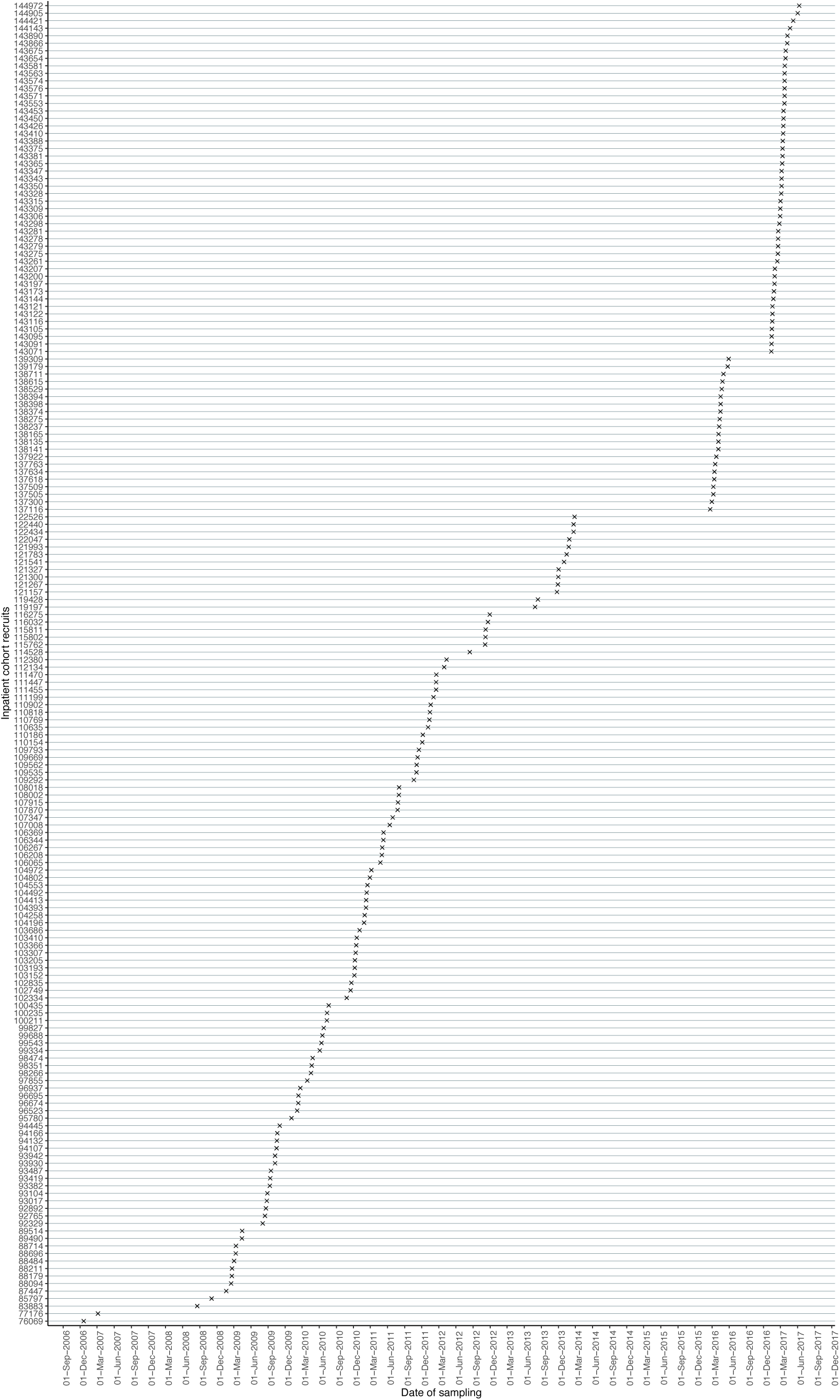
Inpatient cohort comprising of paediatric inpatients admitted to Kilifi County Hospital between 2006 and 2017 with a variety of infectious and non-infectious illnesses. One serological sample was taken at a single time point during admission for medical care. The y-axis contains anonymized codes that are delinked from personally identifiable participant data.

**Supplementary figure 3.**
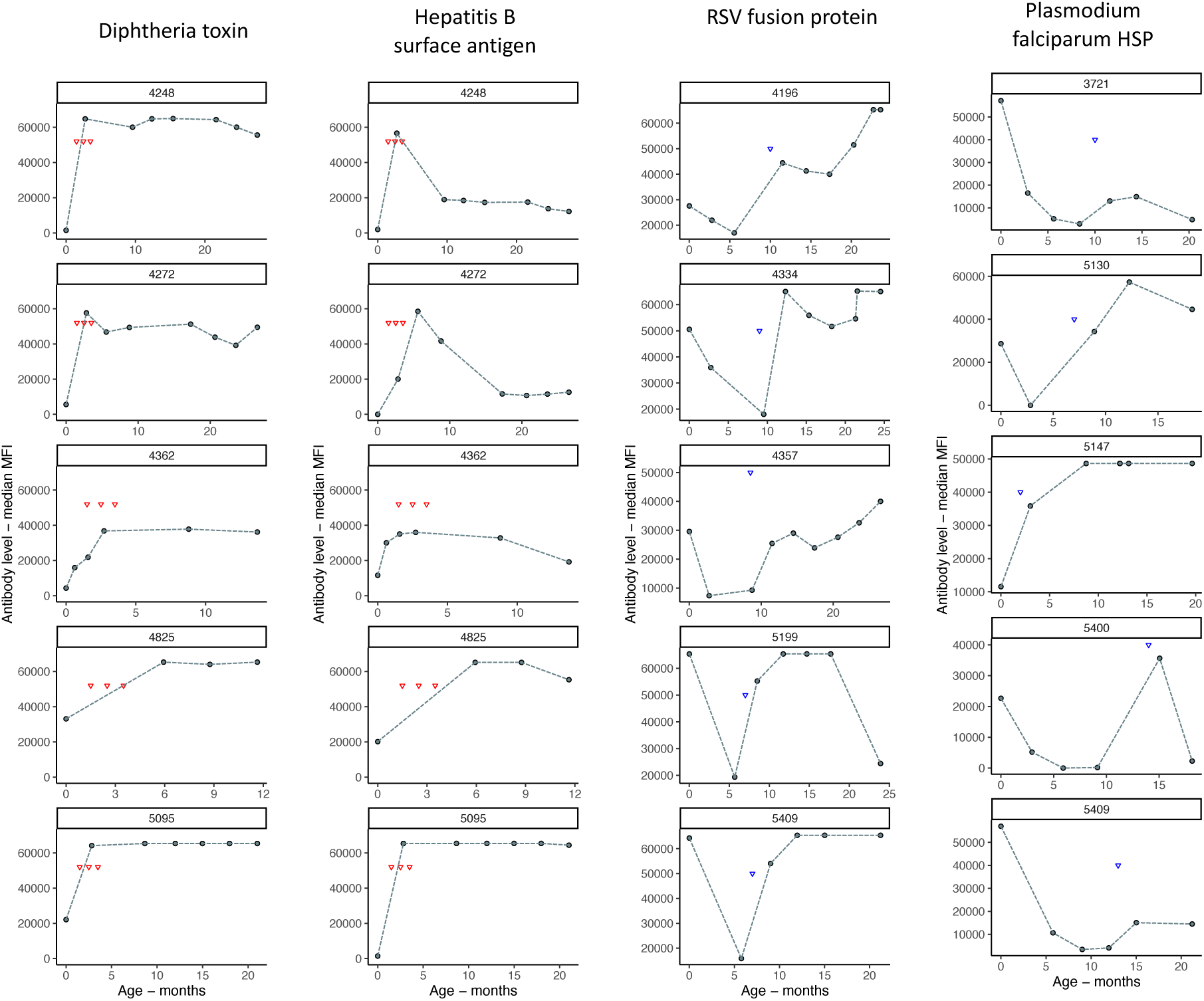
Supplementary figure 3 illustrates our validation approach using diphtheria toxin, hepatitis B surface antigen, RSV fusion protein and plasmodium falciparum HSP. The serological definition of an infection was an increase of ≥ 4000 antibody units (expressed as MFI) between two serial timepoints, and sustained in the subsequent two time points. Each red triangle correlates to the timing of each dose of pentavalent vaccination. The purple triangle indicates the timepoint at which natural infection occurred. Panel headings contain anonymized codes that are delinked from personally identifiable participant data.

**Supplementary figure 4.**
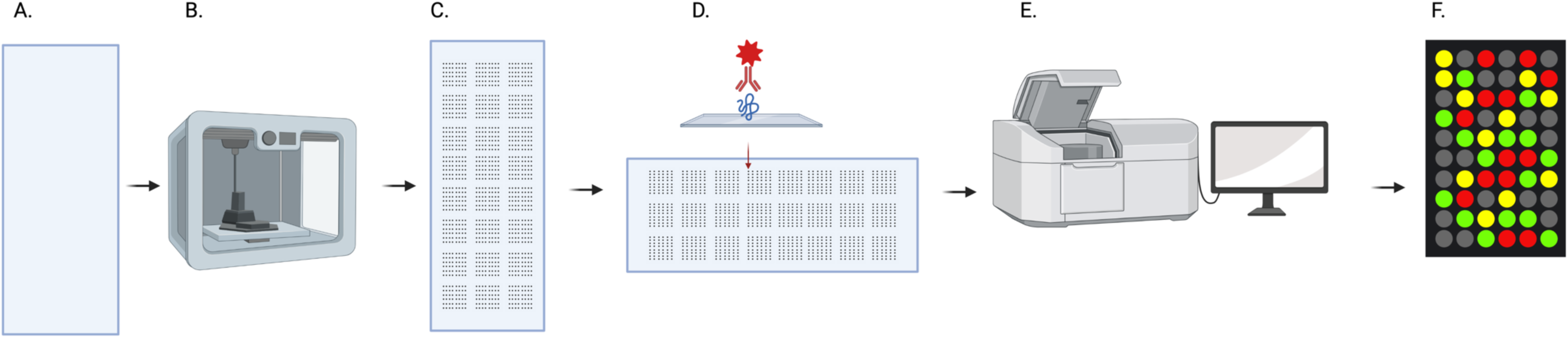
The design of the microarray chip and the analytical sample workflow.

## Supplementary table

**Supplementary table 1.** The complete set of pathogens, strains and antigens that were tested on this platform are listed above. Pathogen-specific antibodies were measured using a customized protein microarray designed to quantify serum IgG to a range of common and emerging infectious diseases.

| ID | Pathogen | Strain/subtype | Antigen | Antigen Code |
| --- | --- | --- | --- | --- |
| 1 | Astrovirus | Type 1 strain A88/2 | Extract | ASTROVT1 |
| 2 | Bordetella pertussis |  | Pertactin | BORDPERT |
| 3 | Chikungunya virus |  | E1 | CCHIKVE1 |
| 4 | Chlamydia pneumoniae |  | PmpD | CHLAMPNEU |
| 5 | Clostridium difficile |  | GDH | CLOSTDEF |
| 6 | Corynebacterium diphtheriae |  | Diphtheria toxin | DIPHTXN |
| 7 | Coxsackie B | B1 | VP1 | COXSACB1 |
| 8 | Cytomegalovirus |  | PP52 | CMVPP52 |
| 9 | Dengue virus | Type 1 | NS1 | DENG1NS1 |
|  | Dengue virus | Type 3 | NS1 | DENG3NS1 |
|  | Dengue virus | Type 4 | NS1 | DENG4NS1 |
| 10 | Ebola virus | Zaire strain | NP | EBOVZAIRE |
| 11 | Enterovirus 71 |  | Toxin | EV71ETOX |
| 12 | Epstein-Barr virus |  | Nuclear antigen 1 | EBV |
| 13 | Helicobacter pylori |  | Hp bacterial antigen | HPYLOR |
| 14 | Hepatitis A virus |  | HAVCR2 | HEPAV |
| 15 | Hepatitis B virus |  | Surface antigen AD | HEPBSAGADR |
| 16 | Herpes simplex virus | Type 1 | Extract | HSV1EXT |
| 17 | Influenza A | A/WSN/1933 | Haemagglutinin | H1N1_HA.1933 |
|  | Influenza A | A/New Caledonia/20/1999 | Haemagglutinin | H1N1_HA.1999 |
|  | Influenza A | A/California/07/2009 | Haemagglutinin | H1N1_HA.2009 |
|  | Influenza A | A/USSR/90/1977 | Haemagglutinin | H1N1_HA.1977 |
|  | Influenza A | A/Panama/2007/1999 | Haemagglutinin | H3N2.1999 |
|  | Influenza A | A/Solomon Islands/3/2006 | Haemagglutinin | H1N1_HA.2006 |
|  | Influenza A | A/Brevig Mission/1/1918 | Haemagglutinin | H1N1_HA.1918 |
|  | Influenza A | A/Puerto Rico/8/1934 | Haemagglutinin | H1N1_HA.1934 |
| 18 | Klebsiella pneumoniae |  | oMPA | KLEBPNEU |
| 19 | Leishmania donovani |  | AdoMetDC | LEISHDON |
| 20 | Mumps virus | Enders | Extract | MUMPSV |
| 21 | Mycoplasma pneumoniae | FH | Extract | MYCPNEU2 |
| 22 | Ngari virus |  | Extract | NGARIV |
| 23 | Norovirus | Type 2 | VLP | NOROVGRP2 |
| 24 | Parvovirus | B19 | Antigen 2 | PARVO1 |
| 25 | Plasmodium falciparum | 3D7 | HSP | PFALHSP |
| 26 | Respiratory syncytial virus | Group A | Fusion protein | RSVAF |
| 27 | Rift valley fever virus |  | Glycoprotein | RVFGC |
| 28 | Rotavirus | P6 | VP6 | ROTAVP6 |
| 29 | Salmonella enterica | Serovar Typhi | H protein | STYPHH |
| 30 | Staphylococcus aureus |  | Alpha Toxin | SAURENTOXA |
| 31 | Streptococcus pneumoniae |  | Pneumolysin | SPNEUMO |
| 32 | Tick-borne encephalitis virus |  | NS1 | TBENS1 |
| 33 | Toxoplasma gondii |  | H4 | TOXOPGON |
| 34 | Usutu virus |  | NS1 | USUVNS1 |
| 35 | Varicella zoster virus | Ellen | Extract | VZV |
| 36 | West Nile virus |  | NS1 | WNVNS1 |
| 37 | Yellow fever virus |  | NS1 | YFVNS1A |
| 38 | Zika virus |  | NS1 | ZIKVNS1 |

